# Covid-19 symptom burden, chronic disease, mental health, and executive function: Multi-country evidence from four African countries

**DOI:** 10.64898/2026.02.16.26346431

**Authors:** Leapetswe Malete, Amara Ezeamama, Chelsi Ricketts, Dale Joachim, Maryam Naghibolhosseini, Mohsen Zayernouri, Reginald Ocansey, Rosemary Chizobam Muomah, Dawn Tladi, Joyce Sifa Ndabi

## Abstract

**Background:** Evidence from high-income countries suggests that COVID-19 may adversely affect cognitive functioning, yet population-based data from African countries remain scarce. Understanding how COVID-19 symptom burden, chronic disease, and mental health intersect to shape cognitive outcomes is critical in low-resource settings disproportionately affected by structural and health system constraints.

**Methods:** Cross-sectional data were collected from 3,058 adults (M_age = 27.2 years) in Botswana, Ghana, Nigeria, and Tanzania between April 2020 and November 2022 using the Sonde Health platform. Participants self-reported sociodemographic characteristics, COVID-19 symptoms, chronic disease diagnoses, mental health symptoms, physical activity, and sedentary behavior. Executive function was assessed using the Stroop Color–Word interference score. Multivariable linear regression models estimated adjusted mean differences in executive function associated with COVID-19 symptom burden and chronic disease, controlling for sociodemographic, health, mental health, and behavioral factors. Effect modification by country was evaluated using interaction terms (p < 0.10).

**Results:** Executive function declined with increasing COVID-19 symptom burden, with Stroop scores decreasing from 0.14 among participants reporting no symptoms to 0.07 among those reporting three or more symptoms (p < 0.001). Being symptom-free was associated with better executive function in Ghana (adjusted mean difference = 0.06; 95% CI: 0.00, 0.11) and Nigeria (adjusted mean difference = 0.07; 95% CI: 0.02, 0.12), but not in Botswana or Tanzania. Lower chronic disease burden was associated with better executive function in Nigeria (adjusted mean difference = 0.16; 95% CI: 0.06, 0.26). Higher educational attainment was consistently associated with better executive function across countries.

**Conclusions:** COVID-19 symptom burden and chronic disease were associated with poorer executive function across the four African countries studied, with substantial cross-country variation. Education emerged as a consistent protective factor. These findings highlight the importance of integrated, context-sensitive approaches that address both physical and mental health to support cognitive well-being during and beyond public health crises.

## Introduction

The COVID-19 pandemic has had profound and multifaceted effects on global health, extending beyond acute infection to long-term challenges for both mental health and cognition. While the neuropsychiatric and psychosocial consequences of COVID-19 are increasingly documented globally, evidence from the African continent remains limited despite the region’s unique vulnerabilities. Low- and middle-income countries (LMICs), which account for over 80% of the world’s population, experienced disproportionate burdens during the pandemic due to fragile health systems, pre-existing chronic disease prevalence, and underinvestment in mental health infrastructure (1). These systemic challenges magnified the psychological toll of the pandemic, particularly through service disruptions, economic instability, and social isolation (2,3).

Mental health symptoms, particularly anxiety and depression, have been shown to increase during the COVID-19 pandemic and are themselves associated with impaired attention and executive functioning. As such, mental health may represent a parallel pathway through which pandemic-related stressors influence cognitive performance, rather than a simple confounder. Across Africa, population-based studies revealed substantial increases in anxiety, depression, and psychological distress during the pandemic (2,4). Risk of anxiety, depression, and psychological distress across Africa during COVID-19 was higher among females, those of lower socioeconomic status, those with greater symptom burden, and individuals with pre-existing chronic conditions, while protective influences such as education, family support, and self-efficacy offered resilience (5). Yet, the delivery of mental health care faced persistent barriers, including stigma, under-resourced systems, and limited digital literacy that constrained alternatives such as telehealth leading many to rely on informal, community-based coping strategies, which, while culturally relevant, were insufficient to address widespread need. Importantly, anxiety and depressive symptoms are not only adverse outcomes of the pandemic but are also known to impair attention, working memory, and executive control, suggesting that mental health may represent a parallel pathway through which COVID-19–related stressors influence cognitive functioning (1,2).

In addition to mental health outcomes, attention has turned to COVID-19’s impact on cognition, particularly cognitive function, which broadly refers to the set of mental processes involved in acquiring knowledge and understanding through thought, experience, and the senses, including attention, memory, processing speed, and reasoning (6). Within this domain, executive function is of particular importance. Executive functions are higher-order cognitive processes, such as inhibitory control, working memory, and cognitive flexibility, that enable individuals to plan, adapt, regulate behavior, and solve problems in a goal-directed manner (7,8). These functions are critical for decision-making and daily adaptive functioning, and their impairment has significant implications for individual well-being and public health. Studies examining cognitive functioning during or following COVID-19 infection have found evidence of impairment, primarily in executive functioning, often described as “brain fog” (9). Mechanisms proposed include neuroinflammation, cerebrovascular changes, and psychosocial stressors that disrupt brain–behavior pathways (10).

Evidence from Africa is minimal but would most likely mirror these concerns. In Egypt, a study of COVID-19 survivors found that they were more likely to have impairments in visuo-executive functions, language, and delayed recall (11). The study also reported that individuals presenting with cognitive impairments were more likely to be older and have a low to moderate level of education. Other studies conducted external to Africa have identified being female and unemployed as additional risk factors for worsening cognitive functions related to COVID-19 (12,13). Mental health challenges associated with COVID-19 illness, including persistent fatigue and decreased interest in pleasure and daily activities, may also co-occur with or exacerbate cognitive difficulties (14,15). Conversely higher educational attainment has consistently emerged as a protective factor, buffering against both mental health declines and cognitive impairment (5).

Importantly, cognitive outcomes are shaped by broader health and behavioral dynamics. Lockdown restrictions contributed to declines in time spent in moderate-to-vigorous-intensity physical activity (MVPA) per week and increases in sedentary behavior (e.g., sitting time), both of which are linked to poorer mental health and potentially reduced cognitive resilience (16,17). Chronic conditions such as hypertension, diabetes, and immunocompromising illnesses, common in many African settings, may further exacerbate vulnerability to cognitive impairment when coupled with COVID-19 symptom burden (18–20). Studies have also shown that patients experiencing COVID-19 symptoms and related cognitive impairments report significantly lower self-rated health compared to those who fully recovered (21). Self-rated health is considered a stronger predictor of future health outcomes and risk of morbidity than clinical measures or laboratory tests, highlighting the value of individual’s own assessments in understanding COVID-19-related impacts (22). Collectively, evidence suggests that cognitive outcomes related to COVID-19 are shaped by a combination of sociodemographic, health, and behavioral factors, highlighting the importance of targeted research on risk and protective mechanisms, particularly given the ongoing and long-term impacts of COVID-19 worldwide (23).

Despite growing global evidence linking COVID-19 to adverse mental health and cognitive outcomes, empirical research from the African continent remains limited. Existing studies are often single-country, hospital-based, or focused primarily on psychological symptoms, restricting understanding of how COVID-19 symptom burden, chronic disease, and mental health intersect to influence cognitive functioning at the population level. Addressing these gaps is particularly important in low- and middle-income countries, where pandemic-related stressors co-occur with structural inequities, constrained health systems, and a high burden of chronic conditions. Hence, the purpose of this study was to examine associations among COVID-19 symptom burden, chronic disease burden, and executive function among adults across four African countries: Botswana, Ghana, Nigeria, and Tanzania, while accounting for mental health, sociodemographic, and behavioral factors. These countries were selected to capture diversity in sociocultural contexts, educational systems, chronic disease profiles, and pandemic responses, enabling cross-country comparisons and the assessment of contextual heterogeneity.

### Research questions and hypotheses

The following research questions and hypotheses were investigated:

1. **Research question 1.** What is the association between COVID-19 symptom burden and executive function after adjusting for sociodemographic characteristics, mental health, self-rated health, and behavioral factors (MVPA and sedentary time)? **Hypothesis 1.** Greater COVID-19 symptom burden will be associated with poorer performance on the Stroop Color-Word Interference Test after adjustment for covariates.
2. **Research question 2.** What is the association between chronic disease burden and executive function after accounting for sociodemographic characteristics, mental health, self-rated health, and behavioral factors (MVPA and sedentary time)? **Hypothesis 2.** A greater chronic disease burden will be associated with poorer Stroop performance after adjustment for covariates.
3. **Research question 3.** Do associations between COVID-19 symptom burden, chronic disease burden, and executive function vary by country and selected demographic characteristics? **Hypothesis 3.** Associations between COVID-19 symptom burden and executive function will differ by country and age, reflecting heterogeneity in contextual, structural, and population-level factors across Botswana, Ghana, Nigeria, and Tanzania.

## Materials and methods

### Participants and procedures

This cross-sectional study is part of a larger project examining physical activity, executive function, and mental health during the COVID-19 pandemic across Botswana, Ghana, Nigeria, and Tanzania. Some data from this project have been previously published, including analyses of age and gender differences in physical activity and depression during COVID-19 in Nigeria (24). The present study addresses distinct research questions focused on executive function and its associations with COVID-19 symptom burden, chronic disease, and mental health.

Ethical approval was obtained from the Institutional Review Boards of the University of Nigeria (UNN/STRACEP/VC/20/2; approved April 27, 2020), University of Botswana (HPDME: 13/18/1; approved September 4, 2020), University of Ghana (ECH162/19-20; approved June 5, 2020), University of Dar es Salaam (SoED-2006; approved June 1, 2020), and Michigan State University (STUDY00004515; approved August 21, 2020). All procedures complied with institutional and national ethical guidelines and the principles of the Declaration of Helsinki. Data were collected between April 2020 and November 2022, with country-specific timelines reflecting institutional ethical approvals, continuing review, and site activation. Data collection at each site was conducted only during periods covered by active ethical approval.

A prior power analysis using G*Power (version 3.1) indicated that a minimum sample size of 250 participants per country would provide adequate statistical power (power = 0.80, *α* = 0.05, effect size: *f^2^*= 0.02) to detect small associations in multivariable regression analyses (25,26). Larger samples were recruited to account for potential missing data and enhance representativeness across settings.

Following ethical approval, trained research assistants recruited participants via email and social media platforms (e.g., WhatsApp). Eligible participants were adults aged 18 years or older. Participants accessed a secure web-based survey hosted on the Sonde Health platform using personal digital devices (e.g., smartphones, tablets, or computers). The survey included an electronic informed consent process that described the study’s purpose, voluntary participation, confidentiality protections, and participants’ rights. Consenting participants completed standardized measures assessing sociodemographic characteristics, COVID-19 symptoms, chronic disease diagnoses, physical activity, sedentary behavior, executive function, and mental health symptoms.

Data were transmitted and stored securely on the Sonde Health cloud platform. Only de-identified data were accessible to investigators for analysis. No personally identifying information was available to the authors during or after data collection, and all analyses were conducted using anonymized datasets.

### Measures

#### Demographic questionnaire

Demographic data were collected on participants’ age, sex, race/ethnicity, area of residence, education level, and employment status. Participants also provided information on COVID-19-related factors, including symptom presentation, COVID-19 testing, and vaccination status at the time of the study. The COVID-19 symptoms assessed included persistent cough, unusual fatigue, loss of taste or smell, skipping meals, unusual headache, running nose, and sore throat. Symptom scores were coded as present (1) or absent (0) and summed to create a total symptom count ranging from 0 to 7. Additional information was collected on participants’ chronic disease diagnoses and health status. Self-reported health status was rated as excellent, very good, good, fair, or unknown, and then dichotomized into *good* (comprising good, very good, and excellent) and *poor* (comprising fair and poor), while excluding participants who reported an unknown rating (27).

#### Physical activity

The IPAQ-SF is a 7-item measure of time spent in MVPA and walking over the past 7 days (28). The IPAQ-SF categories can be analyzed separately to obtain the weekly time (minutes) spent in each activity or used to determine the metabolic equivalent minutes spent in each per week. In this study, physical activity was assessed as time (minutes) spent in MVPA per week to align with WHO (3) guidelines on recommended activity levels. Time (minutes) spent in MVPA per week was calculated by summing the moderate and vigorous intensity PA categories. The IPAQ-SF is shown to have good psychometric properties (28,29). The internal consistency (ω) of scores produced by the IPAQ-SF in this study was 0.50 and considered acceptable (30).

#### Sedentary behavior

Sedentary time was assessed using a single item from the IPAQ-SF, which asks participants to report the duration (minutes per day) of time spent sitting on a weekday over the last 7 days (i.e., During the last 7 days, how much time did you spend sitting on a weekday?) (28). Sitting time is a commonly used and accepted indicator of sedentary time during waking hours (31). Scores produced by the sitting time item on the IPAQ-SF have shown evidence of test-retest reliability (e.g., Spearman correlation coefficients > 0.70) and acceptable criterion validity against accelerometers (32). As per guidelines, data on sitting time (minutes/per day) were reported as median values and interquartile range (IQR) (29).

#### Executive function

Executive function was assessed as the primary outcome using the Stroop Color–Word Test, a widely used neuropsychological measure of cognitive control and inhibitory processing (33). The task was self-administered online in English via a Sonde Health survey platform and consisted of three sequential conditions: (a) a word condition, in which participants read color names (e.g., *red*, *blue*, *green*) printed in black ink; (b) a color condition, in which participants named the ink color of colored rectangles; and (c) a color–word interference condition, in which color names were presented in incongruent ink colors (e.g., the word *red* printed in blue ink), requiring participants to inhibit the automatic reading response and instead identify the ink color. Consistent with prior research, executive function was operationalized using the Stroop Color–Word interference score, analyzed as a continuous measure, and calculated as the difference in performance (response time or errors) between the interference condition and performance predicted from the baseline word and color conditions (34). Higher scores indicate better executive function, reflecting more efficient inhibitory control, cognitive flexibility, and sustained attention under conditions of cognitive conflict.

Because online cognitive tasks can be sensitive to device characteristics (e.g., screen size, input modality) and network-related timing variability, findings are interpreted as associations with performance on this operational measure rather than as definitive clinical assessments of cognitive impairment across settings. Country-level differences in scores may therefore reflect both contextual factors and measurement context (e.g., digital familiarity and testing environment).

#### Anxiety symptoms

The Generalized Anxiety Disorder (GAD-7) scale is a self-report screening tool and severity measure for anxiety (35). The seven items on the GAD-7 ask participants to rate how frequently they have experienced symptoms of anxiety (e.g., excessive worry, nervousness) over the past two weeks. Ratings are provided using a 4-point scale ranging from 0 *(not at all)* to 3 *(nearly every day)*. The GAD-7 produces continuous scores from 0 – 21, which can also be categorized as follows: minimal anxiety (0–4), mild anxiety (5–9), moderate anxiety (10–14), and severe anxiety (15–21). Only continuous scores were examined in this study, with higher scores indicating greater anxiety symptoms. Scores derived from the GAD-7 have shown evidence of reliability and validity in the general population. The internal consistency reliability (ω) of scores produced by the GAD-7 in this study was 0.88.

#### Depressive symptoms

The Patient Health Questionnaire (PHQ-9) is a self-report measure of depressive symptoms and severity (36). Participants are asked to rate how frequently they have experienced symptoms of depression (e.g., little interest in doing things, feelings of hopelessness) over the past 2 weeks. The nine items on the PHQ-9 are scored on a 4-point scale ranging from 0 *(not at all)* to 3 *(nearly every day)*. The PHQ-9 produces continuous scores from 0 – 27, which can also be categorized as follows: minimal depression (0–4), mild depression (5–9), moderate depression (10–14), moderately severe depression (15–19), and severe depression (20–27). Only continuous scores were analyzed in this study, with higher scores indicating greater depressive symptoms. The PHQ-9 scores have good psychometric properties. The internal consistency reliability (ω) of scores produced by the PHQ-9 in this study was 0.89.

### Statistical analysis

Descriptive statistics were calculated for the overall sample, by country, and by COVID-19 symptom burden. Group differences in sociodemographic characteristics, mental health indicators, and health behaviors were assessed using analyses of variance (ANOVA) for continuous variables and chi-square tests for categorical variables. Multivariable linear regression models estimated adjusted mean differences in executive function, measured by Stroop Color–Word interference scores, in relation to COVID-19 and chronic disease symptom burdens.

Covariates were selected *a priori* based on plausible confounding structures and included age, sex, education, employment status, self-rated health, moderate-to-vigorous physical activity (MVPA), and sedentary time; country was included to account for structural differences across settings. Because anxiety (GAD-7) and depressive symptoms (PHQ-9) may function as correlates or potential intermediates linking symptom burden to cognitive performance, models including these variables are interpreted as associations net of concurrent mental health symptomatology, rather than as estimates of causal direct effects.

Potential effect modification by country and selected sociodemographic factors was evaluated by adding two-way interaction terms (e.g., country × COVID-19 symptom burden) to adjusted models. Given the exploratory nature of interaction testing, *p* < 0.10 was used to indicate potential heterogeneity. Analyses were conducted in SAS version 9.4 (SAS Institute Inc., Cary, NC) using two-sided tests with α = 0.05. Models used complete-case observations, and sample sizes varied across analyses due to item non-response.

## Results

### 1. Sample characteristics of the study population

The analytic sample comprised 3,058 adults from Botswana (24.4%), Ghana (28.4%), Nigeria (31.1%), and Tanzania (16.1%), with more than 95% identifying as Black/African descent. Females represented 46.7% of participants, ranging from 37.3% in Ghana to 60.4% in Botswana. The mean age was 27.2 years (SD = 9.2), with the youngest mean age in Ghana (23.6 years) and the oldest in Tanzania (29.4 years). Educational attainment varied across countries: a bachelor’s degree or higher was most common among participants in Tanzania (70.6%) and least common in Botswana (41.6%). Marked country differences were also observed in self-rated health and mental health symptoms, with Botswana reporting the highest anxiety (GAD-7 = 5.92) and depressive symptoms (PHQ-9 = 8.27), and Tanzania the lowest. Mean Stroop Color–Word interference scores ranged from 0.03 in Botswana to 0.24 in Nigeria, while time spent in MVPA was highest in Tanzania and lowest in Ghana (Table 1).

**Table 1.** Sociodemographic, health, and behavioral characteristics of participants from Botswana, Ghana, Nigeria, and Tanzania (N = 3,058).

|  | Overall | Country |  |  |  | P-value |
| --- | --- | --- | --- | --- | --- | --- |
|  | N=3058, 100% | Botswana<br>N=743 (24.4%) | Ghana<br>(N=866, 28.4%) | Nigeria<br>(N=951, 31.1%) | Tanzania<br>(N=491, 16.1%) | Chi-Square<br>/ ANOVA |
|  | n(%)/Mean (SD) | n(%)/Mean (SD) | n(%)/Mean (SD) | n(%)/Mean (SD) | n(%)/Mean (SD) |  |
| **Sex** |  |  |  |  |  |  |
| Female | 1461 (46.72) | 447 (60.41) | 322 (37.27) | 473 (49.79) | 181 (37.24) | <.0001 |
| Male | 1612 (51.55) | 277 (36.43) | 524 (60.65) | 468 (49.26) | 296 (60.91) |  |
| Prefer Not to Say | 54 (1.73) | 16 (2.16) | 18 (2.08) | 9 (0.95) | 9 (1.85) |  |
| **Age (in years)** |  |  |  |  |  |  |
| (Mean, SD) | 27.2 (9.2) | 26.25 (8.79) | 23.58 (5.76) | 30.04 (11.23) | 29.40 (8.06) | <.0001 |
| <20 | 562 (16.97) | 100 (13.46) | 139 (16.05) | 103 (10.83) | 35 (7.12) | <.0001 |
| 20-24 | 1331 (40.19) | 354 (47.64) | 510 (58.89) | 286 (30.07) | 150 (30.55) |  |
| 25-29 | 633 (19.11) | 124 (16.69) | 128 (14.78) | 259 (27.23) | 100 (20.37) |  |
| 30+ | 786 (23.73) | 165 (22.21) | 89 (10.28) | 303 (31.86) | 206 (41.96) |  |
| **Race/Ethnicity** |  |  |  |  |  |  |
| Black/African Descent | 2986 (95.61) | 704 (95.26) | 845 (97.69) | 901 (95.14) | 473 (97.13) | 0.0088 |
| White/European Descent | 31 (0.99) | 3 (0.41) | 4 (0.46) | 14 (1.48) | 1 (0.21) |  |
| Asian/Asian Descent | 79 (2.53) | 24 (3.25) | 13 (1.50) | 24 (2.53) | 12 (2.46) |  |
| Mixed Race | 27 (0.86) | 8 (1.08) | 3 (0.35) | 8 (0.84) | 1 (0.21) |  |
| **Formal Education Completed** |  |  |  |  |  |  |
| Less Than High School Completed | 890 (28.5) | 266 (35.95) | 272 (31.48) | 273 (28.92) | 53 (10.91) | <.0001 |
| Vocational/Technical School | 495 (15.9) | 166 (22.43) | 83 (9.61) | 142 (15.04) | 90 (18.52) |  |
| Bachelor's or Higher | 1735 (55.61) | 308 (41.62) | 509 (58.91) | 529 (56.04) | 343 (70.58) |  |
| **Self-Rated Health** |  |  |  |  |  |  |
| Mean (SD) | 1.78 (0.98) | 1.28 (1.02) | 1.82 (0.93) | 2.06 (0.89) | 1.93 (0.90) | <.0001 |
| Fair/Poor | 420 (13.5) | 197 (27.48) | 81 (9.67) | 47 (4.98) | 31 (6.44) | <.0001 |
| Good | 781 (25.0) | 228 (31.80) | 210 (25.06) | 204 (21.61) | 121 (25.16) |  |
| Very Good | 1061 (34.0) | 186 (25.94) | 330 (39.38) | 336 (35.59) | 179 (37.21) |  |
| Excellent | 860 (27.5) | 106 (14.78) | 217 (25.89) | 357 (37.82) | 150 (31.19) |  |
| **Employment** |  |  |  |  |  |  |
| Student | 1685 (53.9) | 384 (51.82) | 682 (79.03) | 412 (43.41) | 168 (34.64) | <.0001 |
| Formally employed | 466 (14.9) | 79 (10.66) | 34 (3.94) | 255 (26.87) | 80 (16.49) |  |
| Other | 669 (21.4) | 177 (23.89) | 117 (13.56) | 183 (19.28) | 174 (36.08) |  |
| Unemployed | 304 (9.7) | 101 (13.63) | 30 (3.48) | 99 (10.43) | 62 (12.78) |  |
| **Chronic Disease Diagnoses** |  |  |  |  |  |  |
| Asthma | 106 (3.2) | 47 (6.33) | 32 (3.70) | 12 (1.26) | 11 (2.24) | <.0001 |
| High Blood Pressure | 117 (3.5) | 35 (4.71) | 18 (2.08) | 38 (4.00) | 20 (4.07) | 0.0285 |
| Hypercholesterolemia | 66 (1.9) | 10 (1.35) | 15 (1.73) | 25 (2.63) | 10 (2.04) | 0.2716 |
| Other Heart condition | 44 (1.3) | 13 (1.75) | 9 (1.04) | 11 (1.16) | 8 (1.63) | 0.5555 |
| Respiratory Condition | 64 (1.9) | 26 (3.50) | 12 (1.39) | 17 (1.79) | 7 (1.43) | 0.0115 |
| Cancer | 10 (0.3) | 1 (0.13) | 1 (0.12) | 5 (0.53) | 2 (0.41) | 0.3167 |
| Diabetes | 31 (0.9) | 6 (0.81) | 6 (0.69) | 10 (1.05) | 9 (1.83) | 0.2131 |
| Other Chronic Condition | 65 (2) | 29 (3.90) | 15 (1.73) | 12 (1.26) | 8 (1.63) | 0.0011 |
| **Total Chronic Conditions** |  | 0.22 (0.51) | 0.13 (0.39) | 0.14 (0.46) | 0.15 (0.57) | 0.0001 |
| 0 | 2904 (87.68) | 601 (80.89) | 771 (89.03) | 850 (89.38) | 438 (89.21) | <.0001 |
| 1 | 345 (10.42) | 122 (16.42) | 86 (9.93) | 81 (8.52) | 42 (8.55) |  |
| 2+ | 63 (1.9) | 20 (2.69) | 9 (1.04) | 20 (2.10) | 11 (2.24) |  |
| **Total COVID Symptoms** |  |  |  |  |  |  |
| Mean (SD) | 0.9 (1.2) | 1.17 (1.30) | 0.93 (1.19) | 0.74 (1.12) | 0.88 (1.31) | <.0001 |
| 0 | 1610 (49.9) | 301 (40.51) | 416 (48.04) | 548 (57.62) | 271 (55.19) | <.0001 |
| 1 | 826 (25.6) | 196 (26.38) | 242 (27.94) | 233 (24.50) | 108 (22.00) |  |
| 2 | 428 (13.3) | 129 (17.36) | 124 (14.32) | 89 (9.36) | 60 (12.22) |  |
| 3+ | 365 (11.3) | 117 (15.75) | 84 (9.70) | 81 (8.52) | 52 (10.59) |  |
| **Mental Health Questionnaire** | **Mean (SD)** | **Mean (SD)** | **Mean (SD)** | **Mean (SD)** | **Mean (SD)** |  |
| Generalized Anxiety Disorder (GAD-7) Score | 4.2 (4.7) | 5.92 (5.36) | 4.06 (4.57) | 3.54 (4.35) | 3.20 (4.03) | <.0001 |
| PHQ9 Score | 5.7 (5.9) | 8.27 (6.87) | 5.60 (5.62) | 4.37 (5.30) | 4.56 (4.95) | <.0001 |
| Stroop Color-Word Interference Score | 0.1 (0.2) | 0.03 (0.13) | 0.12 (0.20) | 0.24 (0.30) | 0.07 (0.19) | <.0001 |
| **Minutes of Physical Activity** |  |  |  |  |  |  |
| Mean (SD) | 284.1 (452.0) | 256.04 (410.06) | 205.44 (368.17) | 276.11 (434.37) | 467.38 (607.36) | <.0001 |
| None (MVPA=0) | 902 (28.5) | 215 (29.17) | 310 (36.00) | 242 (25.66) | 104 (21.44) | <.0001 |
| Low (MVPA: 1-200) | 1047 (33.1) | 259 (35.14) | 290 (33.68) | 337 (35.74) | 122 (25.15) |  |
| Moderate+ (MVPA >=200) | 1213 (38.4) | 263 (35.69) | 261 (30.31) | 364 (38.60) | 259 (53.40) |  |
| **Sedentary behavior** |  |  |  |  |  | <0.0001 |
| 0 | 210 (6.9) | 24 (3.3) | 57 (6.6) | 81 (8.6) | 48 (9.9) |  |
| 30 | 436 (14.4) | 100 (13.5) | 95 (11.0) | 180 ((19.1) | 61 (12.6)) |  |
| 60 | 483 (16.0) | 101 (13.7) | 111 (12.9) | 196 (20.8) | 75 (15.5) |  |
| 120 | 903 (29.8) | 218 (29.5) | 254 (29.5) | 287 (30.4) | 144 (29.7) |  |
| 240 | 996 (32.9) | 296 (40.1) | 344 (40.0) | 199 (21.1) | 157 (32.4) |  |

**Notes:** Values are n (%) unless otherwise indicated. Continuous variables are presented as mean (SD). Group differences across countries were evaluated using $\chi^2$ tests for categorical variables and one-way ANOVA for continuous variables.
**Abbreviations:** MVPA = moderate-to-vigorous physical activity; GAD-7 = Generalized Anxiety Disorder-7 scale; PHQ-9 = Patient Health Questionnaire-9; Stroop = Stroop Color–Word Interference Test.

### 2. COVID-19 symptom burden and co-occurring health and mental health characteristics

Half of the participants (49.9%) reported no COVID-19 symptoms, 25.6% reported one, 13.3% two, and 11.3% three or more. Greater symptom burden was associated with younger age, a higher proportion of females, poorer self-rated health, and increased prevalence of chronic conditions. Mental health symptoms rose sharply with symptom burden: mean GAD-7 scores increased from 2.41 among those with no symptoms to 7.82 among those with three or more, and PHQ-9 scores from 3.51 to 10.60 (both *p* < 0.0001). In parallel, mean Stroop performance declined from 0.14 to 0.07 as symptom burden increased, indicating poorer executive function. MVPA did not vary significantly across symptom groups (Table 2).

**Table 2.** Participant characteristics by number of self-reported COVID-19 symptoms.

|  | # of COVID-19 Symptoms |  |  |  | P-value |
| --- | --- | --- | --- | --- | --- |
|  | None | One | Two | Three+ | X-square /<br>ANOVA |
|  | N=1610 (49.9%) | N=826 (25.6%) | N=428 (13.3%) | N=365 (11.3%) |  |
| **Biological Sex** | n(%) /Mean (SD) | n(%) /Mean (SD) | n(%) /Mean (SD) | n(%) /Mean (SD) |  |
| Female | 682 (43.61) | 393 (49.56) | 208 (50.00) | 177 (50.57) | 0.0021 |
| Male | 856 (54.73) | 387 (48.8) | 195 (46.88) | 171 (48.86) |  |
| Prefer Not to Say | 26 (1.66) | 13 (1.64) | 13 (3.13) | 2 (0.57) |  |
| **Age (in years)** |  |  |  |  |  |
| Mean, SD | 27.82 (9.13) | 26.54 (9.04) | 26.42 (9.22) | 26.52 (9.92) | 0.0016 |
| <20 | 218 (13.54) | 147 (17.80) | 64 (15.19) | 53 (14.52) | 0.0001 |
| 20-24 | 614 (38.20) | 342 (41.40) | 196 (45.79) | 175 (47.95) |  |
| 25-29 | 333 (20.68) | 157 (19.01) | 79 (18.46) | 64 (17.53) |  |
| 30+ | 444 (27.58) | 180 (21.79) | 88 (20.56) | 73 (20.00) |  |
| **Race/Ethnicity** |  |  |  |  |  |
| Black/African Descent | 1521 (97.19) | 760 (95.84) | 393 (95.16) | 309 (88.54) | <0.0001 |
| White/European Descent | 4 (0.26) | 5 (0.63) | 7 (1.69) | 15 (4.30) |  |
| Asian Descent | 31 (1.98) | 22 (2.77) | 10 (2.42) | 16 (2.25) |  |
| Mixed Race | 9 (0.58) | 6 (0.76) | 3 (0.73) | 9 (2.58) |  |
| **Country** |  |  |  |  |  |
| Botswana | 301 (19.60) | 196 (25.16) | 129 (32.09) | 117 (35.03) | <.0001 |
| Ghana | 416 (26.08) | 242 (31.07) | 124 (30.85) | 84 (25.15) |  |
| Nigeria | 548 (35.68) | 233 (29.91) | 89 (22.14) | 81 (24.25) |  |
| Tanzania | 271 (17.64) | 108 (13.86) | 60 (14.93) | 52 (15.57) |  |
| **Formal Education Completed** |  |  |  |  |  |
| Less Than High School Completed | 437 (27.96) | 209 (26.42) | 119 (28.67) | 124 (35.63) | 0.0087 |
| Vocational/Technical School | 248 (14.87) | 114 (14.41) | 68 (16.39) | 63 (18.10) |  |
| Bachelor's or Higher | 878 (56.17) | 468 (59.17) | 228 (54.94) | 161 (46.26) |  |
| **Employment** |  |  |  |  |  |
| Student | 785 (50.13) | 452 (56.14) | 250 (60.10) | 198 (56.90) | 0.0002 |
| Formally employed | 250 (15.96) | 97 (12.26) | 63 (15.14) | 55 (15.80) |  |
| Other | 363 (23.18) | 169 (21.37) | 79 (18.99) | 56 (16.09) |  |
| Unemployed | 168 (10.73) | 73 (9.23) | 24 (5.77) | 39 (11.21) |  |
| **Self-Rated Health** |  |  |  |  |  |
| Mean (SD) | 2.04 (0.89) | 1.67 (0.97) | 1.50 (0.99) | 1.27 (1.06) | <.0001 |
| Fair/Poor | 89 (5.75) | 104 (13.40) | 70 (17.59) | 100 (29.50) | <.0001 |
| Good | 318 (20.53) | 228 (29.38) | 134 (33.67) | 101 (29.79) |  |
| Very Good | 591 (38.15) | 266 (34.28) | 120 (30.15) | 83 (24.48) |  |
| Excellent | 551 (35.57) | 178 (22.94) | 74 (18.59) | 55 (16.22) |  |
| **Chronic Disease Diagnoses** |  |  |  |  |  |
| Asthma | 32 (1.99) | 25 (3.03) | 26 (6.07) | 23 (6.30) | <.0001 |
| High Blood Pressure | 49 (3.04) | 33 (4.00) | 17 (3.97) | 17 (4.66) | 0.3674 |
| Hypercholesterolemia | 20 (1.24) | 16 (1.94) | 9 (2.10) | 21 (5.75) | <.0001 |
| Other Heart condition | 12 (0.75) | 10 (1.21) | 11 (2.57) | 11 (3.01) | 0.0008 |
| Respiratory Condition | 13 (0.81) | 18 (2.18) | 9 (2.10) | 24 (6.58) | <.0001 |
| Cancer | 2 (0.12) | 3 (0.36) | 3 (0.70) | 2 (0.55) | 0.1982 |
| Diabetes | 12 (0.75) | 9 (1.09) | 5 (1.17) | 5 (1.37) | 0.6223 |
| Other Chronic Condition | 18 (1.12) | 25 (3.03) | 13 (3.04) | 9 (2.47) | 0.0037 |
| **Total Chronic Conditions** |  |  |  |  |  |
| Mean (SD) | 0.10 (0.37) | 0.17 (0.48) | 0.21 (0.59) | 0.31 (0.63) | <.0001 |
| 0 | 1480 (91.93) | 713 (86.32) | 352 (82.24) | 277 (74.89) | <.0001 |
| 1 | 11 (6.89) | 96 (11.62) | 66 (15.42) | 71 (19.45) |  |
| 2+ | 19 (1.18) | 17 (2.06) | 10 (2.34) | 17 (4.66) |  |
| **Mental Health & Cognition** | **Mean (SD)** | **Mean (SD)** | **Mean (SD)** | **Mean (SD)** |  |
| Generalized Anxiety Disorder (GAD-7) Score | 2.41 (3.60) | 5.09 (4.71) | 6.44 (5.08) | 7.82 (5.22) | <.0001 |
| PHQ9_Total | 3.51 (4.60) | 6.54 (5.75) | 8.33 (6.39) | 10.60 (6.52) | <.0001 |
| Stroop Color-Word Interference Score | 0.14 (0.25) | 0.13 (0.25) | 0.10 (0.21) | 0.07 (0.19) | <.0001 |
| **Minutes of Physical Activity** |  |  |  |  |  |
| Mean (SD) | 292.20 (466.19) | 268.92 (437.39) | 278.66 (442.79) | 289.27 (432.60) | 0.6769 |
| None (MVPA=0) | 437 (27.78) | 234 (28.92) | 124 (29.59) | 107 (29.89) | 0.6372 |
| Low (MVPA: 1-200) | 519 (32.99) | 282 (34.86) | 137 (32.70) | 107 (29.89) |  |
| Moderate+ (MVPA >=200) | 617 (39.22) | 293 (36.22) | 158 (37.71) | 144 (40.22) |  |
| **Sedentary behavior** |  |  |  |  |  |
| 0 | 132 (8.7) | 40 (5.2) | 18 (4.5) | 20 (6.1) | <0.0001 |
| 30 | 238 (15.6) | 113 (15.6) | 46 (11.6) | 39 (11.8) |  |
| 60 | 257 (16.9) | 109 (14.1) | 61 (15.3) | 56 (16.9) |  |
| 120 | 456 (29.9) | 210 (27.1) | 134 (33.7) | 103 (31.0) |  |
| 240 | 440 (28.9) | 303 (39.1) | 139 (34.9) | 114 (34.3) |  |

**Notes:** Participant characteristics are shown by number of self-reported COVID-19 symptoms (0, 1, 2, ≥3). P-values derived from $\chi^2$ tests for categorical variables and ANOVA for continuous variables. Mental health and Stroop scores are presented as mean (SD).
**Abbreviations:** MVPA = moderate-to-vigorous physical activity; GAD-7 = Generalized Anxiety Disorder-7; PHQ-9 = Patient Health Questionnaire-

### 3. Association between covid-19 symptom burden and executive function (research question 1)

Multivariable analyses indicated that lower COVID-19 symptom burden was associated with better executive function after adjustment for sociodemographic characteristics, mental health symptoms, self-rated health, MVPA, and sedentary time. Consistent with the category-based estimates presented in Table 3, continuous modeling indicated that each unit decrease in symptom count was associated with an absolute increase of 0.015 points in Stroop Color–Word interference score (95% CI: 0.006, 0.024). A graded dose–response pattern was evident in pooled analyses, with progressively higher performance among participants reporting two, one, or no symptoms compared with those reporting three or more. Similar monotonic trends were observed within Botswana, Ghana, and Nigeria, although statistical significance varied by country (Table 3; Figure 1). Interaction tests indicated no evidence of heterogeneity across countries (p = 0.201).

**Figure 1.**
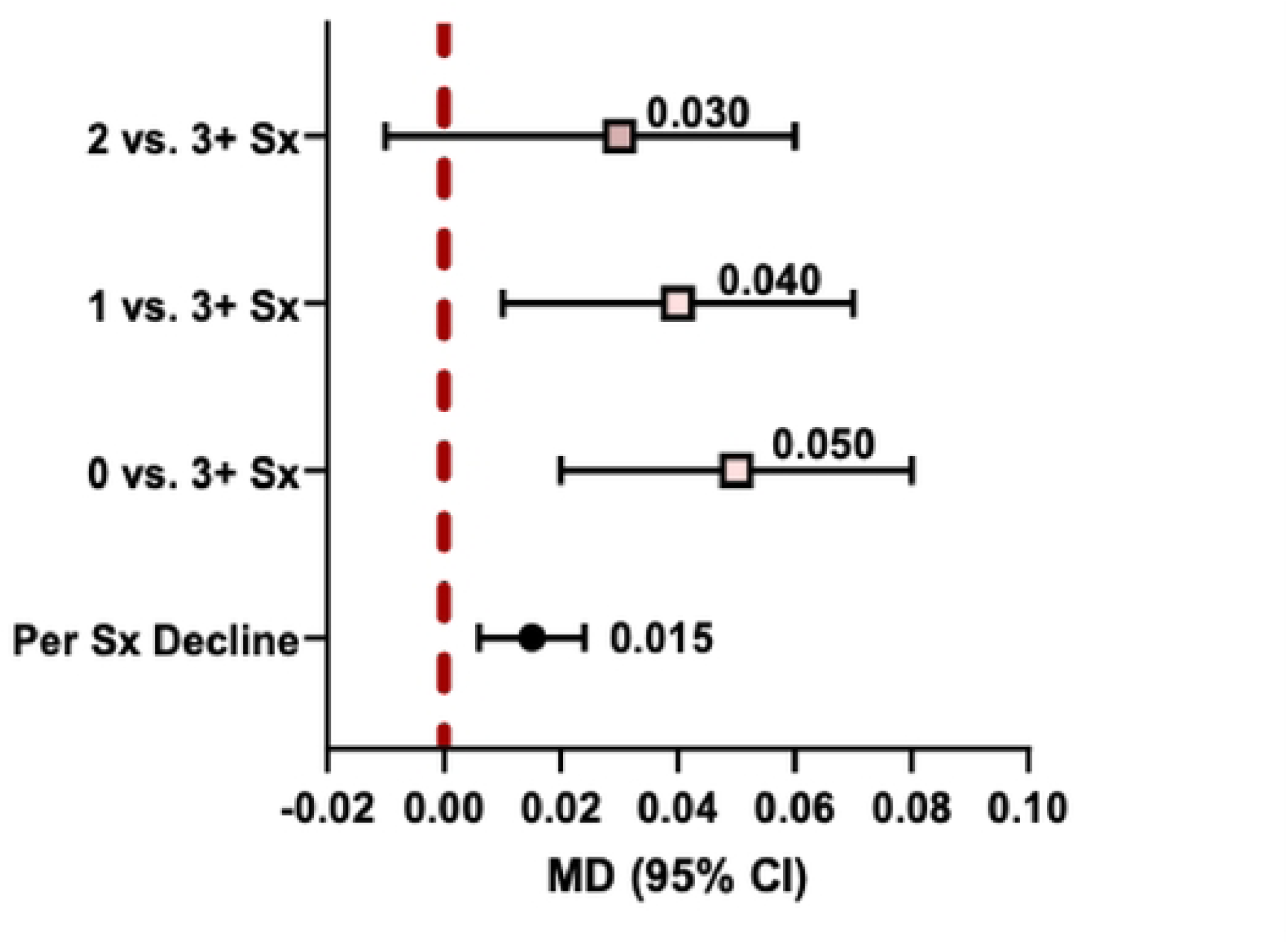
Adjusted mean differences in Stroop Color–Word interference scores by COVID-19 symptom burden. Estimates are adjusted for age, sex, education, employment status, self-rated health, moderate-to-vigorous-intensity physical activity, sedentary time, and mental health symptoms (GAD-7, PHQ-9). Error bars represent 95% confidence intervals.

**Table 3:**
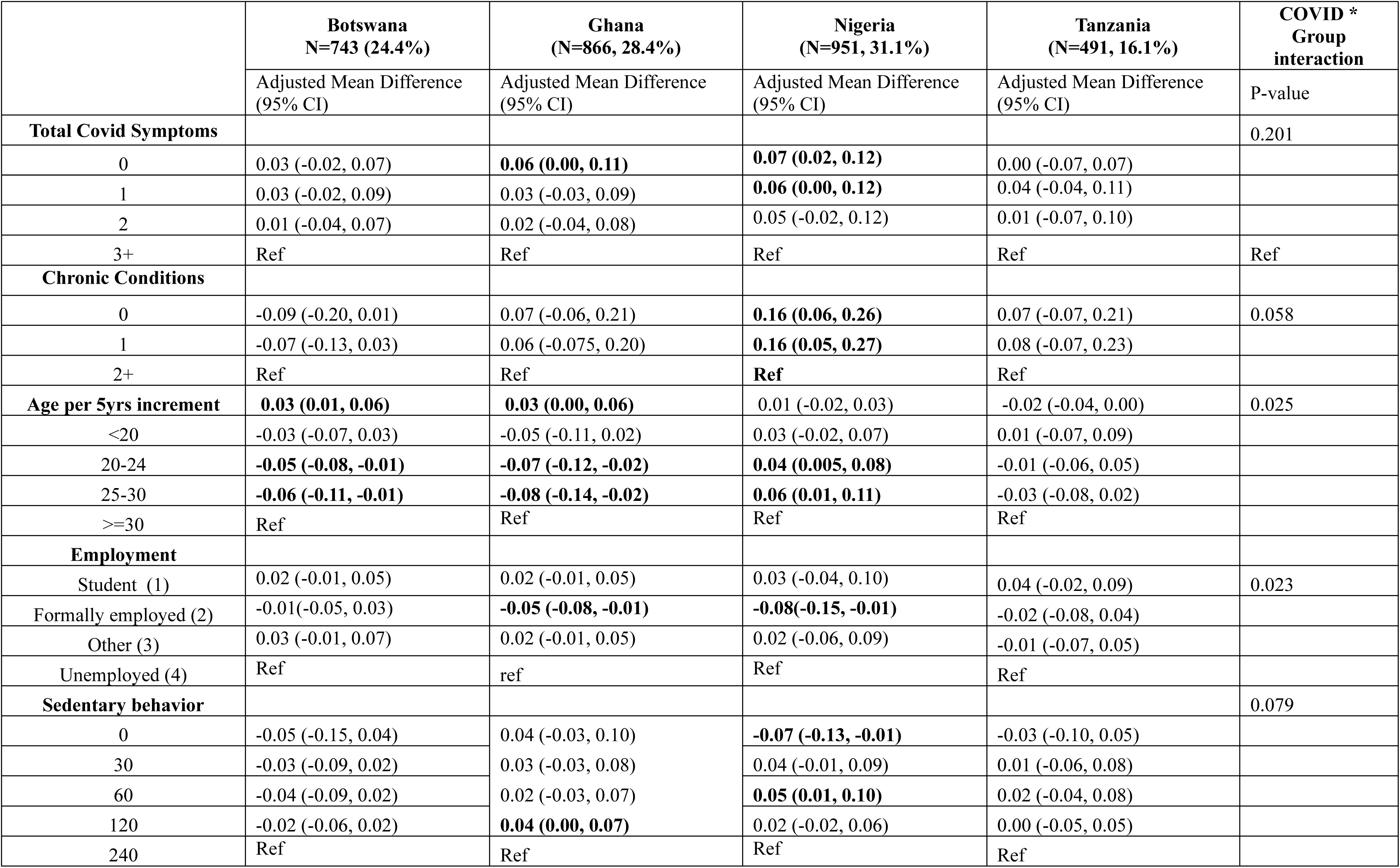
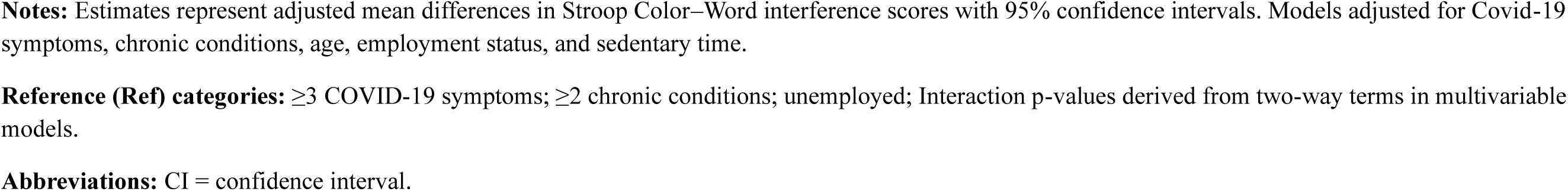
COVID-19, comorbid NCD, and sociodemographic correlates of performance in the Stroop Color-Word interference test among adults from Botswana, Ghana, Nigeria, and Tanzania.

|  | **Botswana**<br>N=743 (24.4%) | **Ghana**<br>(N=866, 28.4%) | **Nigeria**<br>(N=951, 31.1%) | **Tanzania**<br>(N=491, 16.1%) | **COVID \***<br>**Group**<br>**interaction** |
| --- | --- | --- | --- | --- | --- |
|  | Adjusted Mean Difference<br>(95% CI) | Adjusted Mean Difference<br>(95% CI) | Adjusted Mean Difference<br>(95% CI) | Adjusted Mean Difference<br>(95% CI) | P-value |
| **Total Covid Symptoms** |  |  |  |  | 0.201 |
| 0 | 0.03 (-0.02, 0.07) | **0.06 (0.00, 0.11)** | **0.07 (0.02, 0.12)** | 0.00 (-0.07, 0.07) |  |
| 1 | 0.03 (-0.02, 0.09) | 0.03 (-0.03, 0.09) | **0.06 (0.00, 0.12)** | 0.04 (-0.04, 0.11) |  |
| 2 | 0.01 (-0.04, 0.07) | 0.02 (-0.04, 0.08) | 0.05 (-0.02, 0.12) | 0.01 (-0.07, 0.10) |  |
| 3+ | Ref | Ref | Ref | Ref | Ref |
| **Chronic Conditions** |  |  |  |  |  |
| 0 | -0.09 (-0.20, 0.01) | 0.07 (-0.06, 0.21) | **0.16 (0.06, 0.26)** | 0.07 (-0.07, 0.21) | 0.058 |
| 1 | -0.07 (-0.13, 0.03) | 0.06 (-0.075, 0.20) | **0.16 (0.05, 0.27)** | 0.08 (-0.07, 0.23) |  |
| 2+ | Ref | Ref | **Ref** | Ref |  |
| **Age per 5yrs increment** | **0.03 (0.01, 0.06)** | **0.03 (0.00, 0.06)** | 0.01 (-0.02, 0.03) | -0.02 (-0.04, 0.00) | 0.025 |
| <20 | -0.03 (-0.07, 0.03) | -0.05 (-0.11, 0.02) | 0.03 (-0.02, 0.07) | 0.01 (-0.07, 0.09) |  |
| 20-24 | **-0.05 (-0.08, -0.01)** | **-0.07 (-0.12, -0.02)** | **0.04 (0.005, 0.08)** | -0.01 (-0.06, 0.05) |  |
| 25-30 | **-0.06 (-0.11, -0.01)** | **-0.08 (-0.14, -0.02)** | **0.06 (0.01, 0.11)** | -0.03 (-0.08, 0.02) |  |
| >=30 | Ref | Ref | Ref | Ref |  |
| **Employment** |  |  |  |  |  |
| Student (1) | 0.02 (-0.01, 0.05) | 0.02 (-0.01, 0.05) | 0.03 (-0.04, 0.10) | 0.04 (-0.02, 0.09) | 0.023 |
| Formally employed (2) | -0.01(-0.05, 0.03) | **-0.05 (-0.08, -0.01)** | **-0.08(-0.15, -0.01)** | -0.02 (-0.08, 0.04) |  |
| Other (3) | 0.03 (-0.01, 0.07) | 0.02 (-0.01, 0.05) | 0.02 (-0.06, 0.09) | -0.01 (-0.07, 0.05) |  |
| Unemployed (4) | Ref | ref | Ref | Ref |  |
| **Sedentary behavior** |  |  |  |  | 0.079 |
| 0 | -0.05 (-0.15, 0.04) | 0.04 (-0.03, 0.10) | **-0.07 (-0.13, -0.01)** | -0.03 (-0.10, 0.05) |  |
| 30 | -0.03 (-0.09, 0.02) | 0.03 (-0.03, 0.08) | 0.04 (-0.01, 0.09) | 0.01 (-0.06, 0.08) |  |
| 60 | -0.04 (-0.09, 0.02) | 0.02 (-0.03, 0.07) | **0.05 (0.01, 0.10)** | 0.02 (-0.04, 0.08) |  |
| 120 | -0.02 (-0.06, 0.02) | **0.04 (0.00, 0.07)** | 0.02 (-0.02, 0.06) | 0.00 (-0.05, 0.05) |  |
| 240 | Ref | Ref | Ref | Ref |  |

**Notes:** Estimates represent adjusted mean differences in Stroop Color–Word interference scores with 95% confidence intervals. Models adjusted for Covid-19 symptoms, chronic conditions, age, employment status, and sedentary time.
**Reference (Ref) categories:** $\geq 3$ COVID-19 symptoms; $\geq 2$ chronic conditions; unemployed; Interaction p-values derived from two-way terms in multivariable models.
**Abbreviations:** CI = confidence interval.

### 4. Association between chronic disease burden and executive function (research question 2)

Associations between chronic disease burden and executive function varied across countries (Chronic Disease × Country interaction *p* = 0.058). In Nigeria, participants reporting no or one chronic condition exhibited substantially higher Stroop performance compared with those reporting two or more conditions (adjusted mean difference = 0.16, 95% CI: 0.06, 0.26). Directionally similar but non-significant associations were observed in Ghana and Tanzania. In contrast, in Botswana, a qualitatively different pattern emerged: lower chronic disease burden was associated with lower Stroop performance, although the confidence intervals included the null (Table 3).

### 5. Cross-country and demographic variation (research question 3)

#### 5.1 Country-level patterns in executive function

Country-level differences were observed in associations between COVID-19 symptom burden, chronic disease burden, and Stroop performance. While the direction of associations between COVID-19 symptoms and Stroop performance was broadly consistent, effect sizes were larger in Ghana and Nigeria than in Botswana and Tanzania (Table 3).

#### 5.2 Age-specific variation in associations with executive function

Age modified the association between COVID-19 symptom burden and executive function (COVID-19 × Age interaction *p* = 0.028). Among participants aged 25 years and older, lower symptom burden was associated with progressively higher Stroop performance, whereas no clear dose–response pattern was observed among participants younger than 25 years (Figure 2).

**Figure 2.**
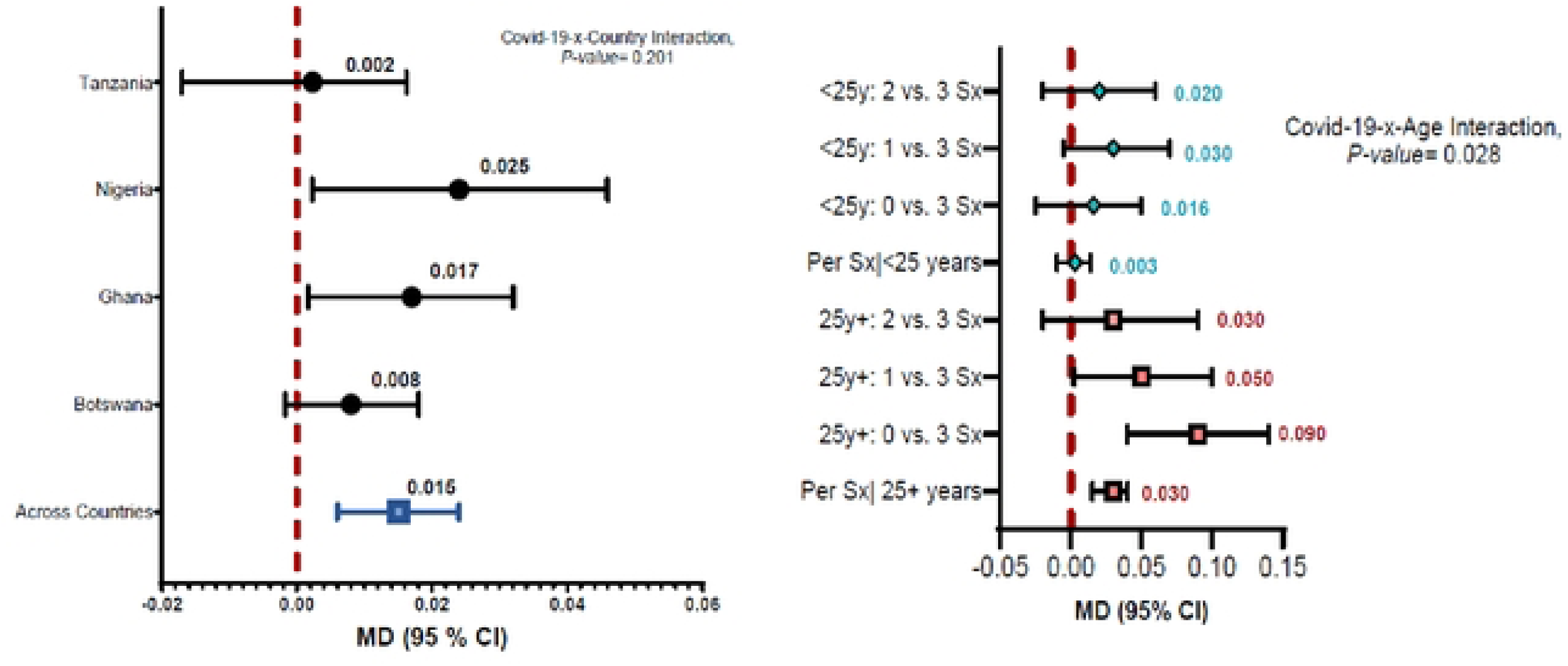
Adjusted mean Stroop Color–Word interference scores by COVID-19 symptom burden, country, and age group. Adjusted mean differences compare participants reporting 0, 1, or 2 symptoms with those reporting ≥3 symptoms within age groups (<25 and ≥25 years). Estimates are adjusted for covariates listed in Table 3. Error bars represent 95% confidence intervals.

#### 5.3 Sociodemographic factors exhibiting country-specific associations

In adjusted models, formal employment was associated with poorer executive function in Ghana (adjusted mean difference = −0.05, 95% CI: −0.08, −0.01) and Nigeria (adjusted mean difference = −0.08, 95% CI: −0.15, −0.01), but not in Botswana or Tanzania. Female sex was associated with modestly higher Stroop performance only in Tanzania (adjusted mean difference = 0.03, 95% CI: 0.00, 0.07). No significant sex associations were observed in Botswana, Ghana, or Nigeria. Sedentary time showed no consistent associations across countries (Table 3). Tests for interaction indicated no evidence of heterogeneity in the overall association across countries (COVID-19 × Country interaction p = 0.201; Figure 2).

### 6. Country-invariant correlates of executive function

Educational attainment was the only country-invariant correlate of executive function across fully adjusted models. Participants with primary education or less exhibited significantly lower Stroop performance compared with those holding a bachelor’s degree or higher (adjusted mean difference = −0.03, 95% CI: −0.05, −0.01). Mental health symptoms, MVPA, and self-rated health were not significantly associated with Stroop performance in fully adjusted models, and no evidence of effect modification by country was observed for these factors (all interaction p-values > 0.25; Table 4).

**Table 4.** Country-invariant associations with Stroop performance across four countries (Botswana, Ghana, Nigeria, Tanzania).

|  | Crude Association |  | Adjusted Association |  | Covid*Group Interaction | Country * Group interaction<br>P-value |
| --- | --- | --- | --- | --- | --- | --- |
|  | LSM (SE) | Difference (95% CI) | LSM (SE) | Difference (95% CI) |  |  |
| **Sex** |  |  |  |  |  |  |
| Prefer not to disclose | 0.045 (0.04) | -0.09 (-0.16, -0.02) | 0.06 (0.03) | -0.05 (-0.12, 0.01) | 0.9485 | 0.5519 |
| Female | 0.12 (0.01) | -0.01 (-0.03, 0.01) | 0.11 (0.01) | -0.01 (-0.02, 0.01) |  |  |
| Male | 0.13 (0.01) | Ref | 0.11 (0.01) | Ref |  |  |
| **Formal Education Completed** |  |  |  |  |  |  |
| <=primary school completed | 0.11 (0.01) | **-0.03 (-0.05, -0.01)** | 0.07 (0.01) | **-0.03 (-0.05, -0.01)** | 0.9880 | 0.7149 |
| Technical/Vocational School | 0.11 (0.01) | **-0.03 (-0.05, -0.01)** | 0.08 (0.02) | -0.02 (-0.04, 0.01) |  |  |
| Bachelors/Higher | 0.14 (0.01) | Reference | 0.10 (0.01) | Ref |  |  |
| **Minutes of Physical Activity** |  |  |  |  |  |  |
| None (0) | 0.12 (0.01) | -0.01 (-0.03, 0.01) | 0.079 (0.01) | -0.01 (-0.03, 0.01) | 0.1379 | 0.9532 |
| Low MVPA (1-200) | 0.12 (0.01) | -0.01 (-0.03, 0.01) | 0.081 (0.014) | -0.01 (-0.04, 0.01) |  |  |
| Moderate+ MVPA (>=200) | 0.13 (0.01) | Reference | 0.092 (0.01) | Ref |  |  |
| **Self-Rated Health** |  |  |  |  |  |  |
| Fair/Poor | 0.08 (0.01) | **-0.06 (-0.09, -0.03)** | 0.10 (0.02) | 0.03 (-0.005, 0.056) | 0.869 | 0.252 |
| Good | 0.11 (0.01) | **-0.03 (-0.06, -0.01)** | 0.08 (0.01) | 0.01 (-0.01, 0.03) |  |  |
| Very Good | 0.14 (0.01) | -0.01 (-0.03, 0.01) | 0.08 (0.01) | 0.01 (-0.01, 0.03) |  |  |
| Excellent | 0.15 (0.01) | Reference | 0.07 (0.01) |  |  |  |

**Notes:** Country-invariant associations estimated from multivariable models including all countries with adjustment for sex, education, moderate -to-vigorous physical activity, and self-rated health. Interaction p-values > .25 indicate no evidence of effect modification by country. **Abbreviations:** LSM = least-squares mean; SE = Standard Error; MVPA = moderate-to-vigorous physical activity; CI = confidence interval.

## Discussion

The purpose of this study was to examine the associations among COVID-19 symptom burden, chronic disease, sociodemographic factors, and executive function across four African countries during the COVID-19 pandemic. Overall, the findings largely supported the study hypotheses. Greater COVID-19 symptom burden and chronic disease comorbidity were associated with poorer executive function, although the magnitude and, in some cases, direction of these associations varied across countries. Education consistently emerged as a protective factor, while physical activity was not associated with Stroop performance. Overall, these findings highlight the importance of shared biological mechanisms, validate the multi-country approach, and underscore the role of context-specific determinants in shaping cognitive health during public health crises. A detailed discussion of our findings is provided in the sections below. These interpretations are based on adjusted mean differences presented in Tables 3 and 4.

### COVID-19 symptom burden and executive function

Consistent with our hypotheses and prior global evidence, higher COVID-19 symptom burden was associated with poorer executive function, as measured by the Stroop Color-Word interference test (Zhou et al., 2020; Hampshire et al., 2022). This pattern aligns with growing evidence that both acute and post-acute COVID-19 symptoms can impair executive functioning and attention. Although biological mechanisms were not directly assessed in this study, plausible pathways include neuroinflammation, immune dysregulation, cerebrovascular injury, and indirect effects related to fatigue, sleep disruption, and psychological stress (14,37). Notably, associations between COVID-19 symptom burden and executive function were strongest in Ghana and Nigeria, suggesting that biological vulnerability may be amplified by sociocultural or systemic factors. Differences in healthcare access, economic disruption, public health messaging, and social stressors may have shaped the extent to which symptom burden translated into cognitive impairment across settings (1,4). These findings underscore the need to interpret pandemic-related cognitive outcomes within broader structural and contextual environments.

### Chronic disease burden and executive function

The observed association between greater chronic disease burden and poorer executive function is consistent with extensive evidence linking cardiometabolic and inflammatory conditions to cognitive decline (38). Chronic conditions may increase susceptibility to cognitive impairment through cumulative inflammatory burden, metabolic dysregulation, and vascular compromise. The divergent pattern observed in Botswana, where lower chronic disease burden was associated with poorer Stroop performance, warrants careful interpretation. One possible explanation is behavioral compensation, whereby individuals with known comorbidities may have adhered more closely to public health guidance or engaged in proactive health-monitoring behaviors during the pandemic. Contextual factors specific to Botswana, including stringent lockdown measures and strong social norms around compliance, may have contributed to this pattern, although these mechanisms were not directly measured in the present study. Alternatively, sampling variation or reporting bias may partially explain these findings. Longitudinal and mixed-method research is needed to clarify whether health awareness, adaptive coping, or unmeasured contextual factors underlie this unexpected association.

### Mental health as a co-occurring pathway

Mental health symptoms emerged as a prominent co-occurring burden, increasing sharply with COVID-19 symptom severity. Anxiety and depressive symptoms are well documented to disrupt attentional control and executive processes, potentially compounding cognitive vulnerability during periods of sustained stress. Importantly, associations between COVID-19 symptom burden and executive function persisted after adjustment for mental health, indicating that cognitive effects were not solely attributable to psychological distress. This pattern supports conceptualizing mental health as a parallel, interacting pathway rather than a complete explanatory mechanism and highlights the need for integrated cognitive and mental health responses during pandemics.

#### Sociodemographic and behavioral influences on executive function

Age-related patterns varied across countries, underscoring the interplay of biological, educational, and sociocultural factors in shaping executive function. In Botswana and Ghana, older participants demonstrated better Stroop performance, potentially reflecting accumulated cognitive reserve or cohort differences in educational attainment (39). In contrast, younger adults in Nigeria performed better, which may relate to faster processing speed, greater digital literacy, or more frequent engagement in cognitively demanding environments. These divergent age patterns highlight the importance of contextualizing cognitive aging within local educational and occupational landscapes.

Employment status also demonstrated context-specific associations. Formal employment was linked to poorer executive function in Ghana and Nigeria, possibly reflecting heightened occupational stress, reduced recovery time following illness, or sustained cognitive load during the pandemic. The absence of similar associations in Botswana and Tanzania may indicate differences in labor conditions, economic security, or workplace adaptations that mitigated cognitive strain. Across all four countries, higher educational attainment was consistently associated with better executive function, reinforcing education’s role in fostering cognitive reserve, metacognitive regulation, and adaptive problem-solving (7). This robust association underscores the long-term cognitive benefits of educational investment. In contrast, physical activity was not associated with Stroop performance, diverging from pre-pandemic literature (40). This null finding may reflect reliance on self-reported activity measures, reduced variability in activity during lockdowns, or the predominance of other health stressors during the pandemic.

### Cross-country variation and contextual determinants

Substantial cross-country heterogeneity in associations underscores the importance of contextually grounded analyses. Although COVID-19 symptom burden and chronic disease were generally associated with poorer cognitive performance, the strength and consistency of these relationships varied across Ghana, Nigeria, Botswana, and Tanzania. These differences likely reflect variation in health system capacity, educational structures, economic resilience, and pandemic response strategies (1). Sociocultural factors may have further shaped these patterns. Differences in health literacy, trust in health institutions, and community support structures can influence symptom reporting, stress exposure, and cognitive resilience (2,4). Measurement context may also have contributed, as variation in digital literacy and familiarity with language-based cognitive assessments could influence performance on the Stroop task. Collectively, these findings reinforce the value of comparative research for understanding how structural and social determinants shape cognitive health disparities across African settings.

### Limitations and future directions

Several limitations should be acknowledged. The cross-sectional design precludes causal inference and limits assessment of temporal ordering among COVID-19 symptom burden, mental health symptoms, and executive function; thus, mental health may represent antecedents, correlates, or consequences of cognitive performance. Recruitment via email and social media and completion of an English-language, self-administered web-based Stroop task likely yielded a sample with greater digital access and educational attainment than the general adult population, which may limit generalizability. Although the Stroop interference score is a widely used index of inhibitory control, online administration can be influenced by device characteristics and network-related timing variability, potentially contributing to cross-country differences independent of neurocognitive function. Reliance on self-reported COVID-19 symptoms, chronic conditions, and physical activity introduces potential recall and classification bias. Finally, the extended data collection period (April 2020–November 2022) spanned distinct pandemic phases that varied across countries; unmeasured time-varying contextual factors may therefore confound observed associations.

Future longitudinal and population-based studies in African settings should validate digital cognitive tasks across languages and devices, including the Stroop platform used here, incorporate clearer indicators of infection timing and long-COVID status, and evaluate whether changes in mental health and cardiometabolic risk mediate cognitive outcomes over time.

### Health and policy implications

These findings have important implications for public health and policy in African contexts. Context-specific strategies are needed to address the cognitive and psychosocial consequences of pandemics. Integrating cognitive screening into primary healthcare, particularly for individuals with high COVID-19 symptom burden or chronic conditions, may facilitate early identification and timely intervention. These priorities highlight the need to invest in culturally valid digital assessment tools and to implement routine cognitive and mental health monitoring within primary care and pandemic-response systems. In addition, sustained investments in education and chronic disease prevention represent long-term strategies to strengthen cognitive resilience and reduce health-related inequities during future public health crises.

## Conclusion

This study provides novel multi-country evidence on the associations between COVID-19 symptom burden, chronic disease, and executive function in four countries representing three African regions (East, West, and Southern Africa). The findings reveal both universal and context-specific determinants of cognitive performance, highlighting the intersecting roles of biological vulnerability, mental health, education, and structural conditions. Education emerged as a consistent protective factor, underscoring the importance of investing in human capital for cognitive and societal resilience during and beyond public health emergencies.

## Data Availability

De-identified data supporting the findings of this study will be available in Figshare at the time of publication: https://doi.org/10.6084/m9.figshare.31306807. The dataset will be shared under a license permitting non-commercial reuse with appropriate citation. Study instruments and analytic code are available from the corresponding author upon reasonable request.

## Acknowledgments

The research team would like to thank the following individuals for their contributions to the project: Vida Korleki Nyawornota, Clement Adamba, JohnBosco C. Chukwuorji, Omphile Hubona, Sampson K. Nwonyi, Doris A. Tay, Oscar C. Nyanyofio, Aarushi Lokhande, Madison Chambers, and Seoah An.

## Supporting information

**S1 Fig. Adjusted mean differences in Stroop Color–Word interference scores by COVID-19 symptom burden.** Estimates are adjusted for age, sex, education, employment status, self-rated health, moderate-to-vigorous-intensity physical activity, sedentary time, and mental health symptoms (GAD-7, PHQ-9). Error bars represent 95% confidence intervals.

**S2 Fig. Adjusted mean Stroop Color–Word interference scores by COVID-19 symptom burden, country, and age group.** Adjusted mean differences compare participants reporting 0, 1, or 2 symptoms with those reporting ≥3 symptoms within age groups (<25 and ≥25 years). Estimates are adjusted for covariates listed in Table 3. Error bars represent 95% confidence intervals.

**S1Table. Sociodemographic, health, and behavioral characteristics of participants from Botswana, Ghana, Nigeria, and Tanzania (N = 3,058).** Values are mean (SD) or n (%). Executive function was measured using the Stroop Color–Word interference score. Mental health symptoms were assessed using GAD-7 and PHQ-9. Group differences were evaluated using ANOVA or chi-square tests.

**Abbreviations:** SD, standard deviation; MVPA, moderate-to-vigorous physical activity; GAD-7, Generalized Anxiety Disorder-7; PHQ-9, Patient Health Questionnaire-9.

**S2 Table. Participant characteristics by number of self-reported COVID-19 symptoms.** Values are mean (SD) or n (%). Executive function was assessed using the Stroop Color–Word interference score. Differences across symptom categories were evaluated using ANOVA or chi-square tests.

**Abbreviations:** SD, standard deviation; MVPA, moderate-to-vigorous physical activity; GAD-7, Generalized Anxiety Disorder-7; PHQ-9, Patient Health Questionnaire-9.

**S3 Table. Adjusted mean differences in Stroop Color–Word interference scores according to COVID-19 symptom burden and chronic conditions, by country.** Estimates are adjusted mean differences from multivariable linear regression models controlling for age, sex, education, employment, self-rated health, MVPA, sedentary time, GAD-7, PHQ-9, and country. Positive values indicate better executive function.

**Abbreviations:** CI, confidence interval; MVPA, moderate-to-vigorous physical activity.

**S4 Table. Country-invariant associations with Stroop Color–Word interference scores across four countries.** Estimates are adjusted mean differences from multivariable linear regression models controlling for sociodemographic, health, behavioral, and mental health factors.

**Abbreviations:** CI, confidence interval; MVPA, moderate-to-vigorous physical activity.

